# Assessment of Gut Microbial β-Glucuronidase and β-Glucosidase Activity in Women with Polycystic Ovary Syndrome

**DOI:** 10.1101/2023.04.06.23288218

**Authors:** Jalpa Patel, Hiral Chaudhary, Bhavin Parekh, Rushikesh Joshi

**Affiliations:** Department of Biochemistry and Forensic Science, University School of Sciences, Gujarat University, Ahmedabad-380009, Gujarat, India; School of Applied Sciences and Technology, Gujarat Technological University, Ahmedabad-382424, Gujarat, India

**Author notes:** **Details of correspondence** Dr. Rushikesh Joshi, Ph.D., Assistant professor, Department of Biochemistry and Forensic Science, University, School of Sciences, Gujarat University, Ahmedabad 380009, Gujarat, India., Dr. Bhavin Parekh, Ph.D., School of Applied Sciences & Technology, Gujarat Technological University, Ahmedabad-382424, Gujarat, India.

**Keywords:** Gut microbiota, β-Glucuronidase, β-Glucosidase, Polycystic ovary syndrome, Stool specimen

## Abstract

**Objective:** To identify Gut microbial β-Glucuronidase and β-Glucosidase activity in polycystic ovary syndrome (PCOS) and reveal a possible correlation between gut bacterial enzyme activities and estrogen.

**Design:** Case-Control Study.

**Subjects:** Reproductive-aged women with PCOS (n=23) and controls (n =25) from the Health Centre of Gujarat University.

**Interventions:** Spectrophotometric analysis of β-Glucuronidase and β-Glucosidase activity of fecal samples from patients and clinical parameters (including body mass index, endocrine hormone levels, and hirsutism) collected for correlation analysis.

**Primary outcome:** Identification of gut bacterial β-Glucuronidase and β-Glucosidase activity differences and clinical parameters.

**Results:** Compared to the controls, PCOS women had considerably higher levels of β-glucuronidase activity, having statistically significant p-value (0.05 ± 0.1vs. 0.04± 0.1; *p=* 0.006). We observed a higher trend of β-glucosidase activity in PCOS women compared to the control (0.13 ± 0.08 vs. 0.09 ± 0.05; *p=* 0.06).

**Conclusion:** We observed a strong trend toward increased levels of β-glucuronidase and β-glucosidase activity in PCOS women compared to healthy control women. This inference requires further validation through studies with a larger sample size. However, if validated, we suggest that β-glucosidase levels can be considered a putative biomarker for PCOS women with metabolic disturbances and might help personalize the treatment.

**Graphical abstract:** 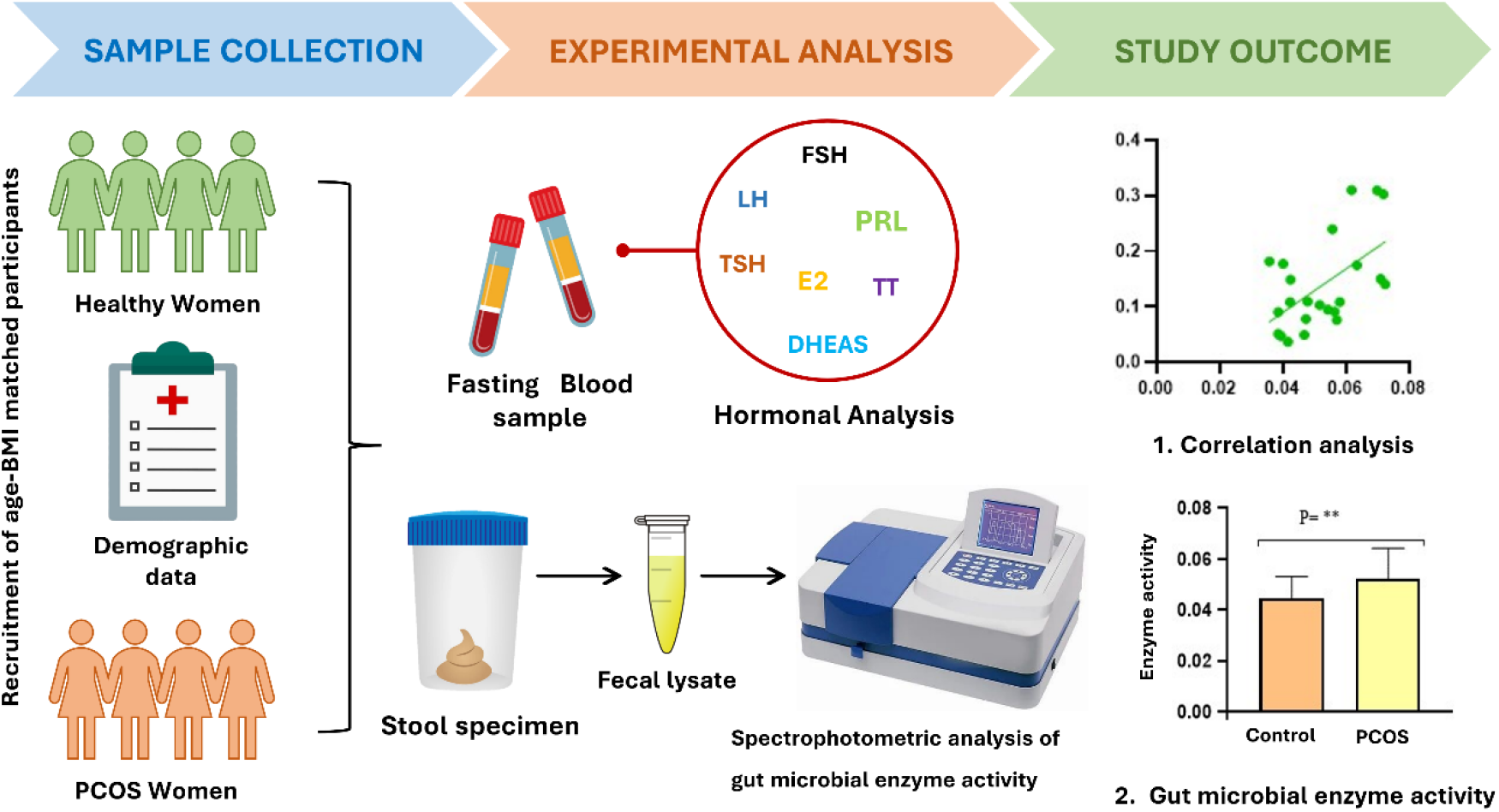

## Introduction

Polycystic ovarian syndrome (PCOS) is a common endocrine condition in women of reproductive age, with an estimated worldwide prevalence of 8% to 13%. PCOS is the leading cause of female infertility (1). PCOS is characterized by polycystic ovaries, chronic anovulation, hyperandrogenism, hyperinsulinemia, abdominal obesity, and dyslipidemia, which can lead to serious long-term issues such as endometrial hyperplasia, type 2 diabetes, and coronary artery disease (2)(3). The pathogenesis of PCOS remains nebulous. However, it is considered multifaceted, involving genetics, the intrauterine environment, and lifestyle choices (4).

The gut microbiota functions as an endocrine organ that can affect other distant organs (5). Using metagenomics tools, researchers have shown that many chronic conditions, such as obesity, diabetes, non-alcoholic fatty liver disease (NAFLD), and PCOS, are all linked to gut microbial dysbiosis (6). Likewise, the role of gut microbes has recently emerged as an exciting research area in PCOS pathology (7). Since 2017, more than a dozen clinical cross-sectional studies have shown that PCOS correlates with gut dysbiosis, while the taxa vary among these studies. Most studies have shown reduced α-diversity and β-diversity, both of which are reported to be associated with endocrine and metabolic abnormalities (8).

Most of these cross-sectional studies have used 16S rRNA gene amplicon sequencing. Although 16S rRNA sequencing provides accurate taxonomic resolution down to the genus level, it cannot measure specific bacterial functions, which may better indicate how bacteria affect PCOS pathology (9). However, PCOS-specific bacterial functions have been scantly explored as characterized by microbial enzymatic activity.

Since gut microbial β-glucuronidase and β-glucosidase enzymes, found in various species of gut bacteria, mediate hormonal (i.e., estrogen) and metabolic homeostasis (10), we wondered whether levels of these enzymes differ between Polycystic ovary syndrome (PCOS) women and healthy women. This is the first study evaluating β-glucuronidase and β-glucosidase activity in stool specimens of women with and without PCOS. The gut microbial enzyme activity was measured in triplicate from the same stool specimen collected independently by each participant. Herein, we measured and correlated the enzyme activities with the clinical parameters.

## MATERIALS AND METHODS

### Participants and Ethics

We recruited forty-eight premenopausal women, of whom twenty-three were PCOS women, and twenty-five were healthy controls. These women, aged 17-42 years, were enrolled from individuals who visited the Health Centre, Gujarat University (Gujarat, India) between December 2019 and December 2022. Diagnosis criteria for PCOS, established by the Health Centre of the Institute, were based on the Rotterdam Criteria and the currently available evidence in Indian women. In our study, to avoid confusion due to the different diagnostic criteria, each participant had met two out of three features based on Rotterdam criteria as follows: Oligo- or anovulation, signs of hyperandrogenism (clinical and biochemical), and polycystic ovaries (PCO, at least 12 follicles measuring 2-9 mm or volume of the ovary > 10 cm3) (Rotterdam ESHRE/ASRM-Sponsored PCOS Consensus Workshop Group, 2004). The controls had no history of diagnosed PCOS.

We recorded multiple clinical parameters, including medical history, dietary habits, and family history. All subjects received a standard physical examination of height, weight, waist circumference, and hip circumference in light clothing without shoes. BMI and waist-to-hip ratio (WHR) were calculated. This study was endorsed by the Ethical Committee of Gujarat University (GU-IEC(NIV)/02/Ph.D./007). All participants provided informed consent before enrolment. All participants were verbally explained about the research and asked to sign the bilingual (English and Gujarati) informed consent form.

### Inclusion and Exclusion criteria

The inclusion criteria for this study were (a) 17-45 years of age and (b) Indigenous ethnicity. In addition, individuals with one of the following conditions have been excluded from the study: pregnancy, hypertension, smoking, thyroid dysfunction, Cushing’s syndrome, adrenal disorders, hyperprolactinemia, gastrointestinal disease, and Diabetes mellitus. Participants also had no administration of hormonal medication, an insulin sensitizer, or antibiotics within the preceding three months.

### Laboratory Measurements

Baseline fasting blood samples were taken on 2-5 days of the menstrual cycle for hormonal and metabolic measurements. The serum aliquots have been stored at -80°C until analysis. We measured levels of follicle-stimulating hormone (FSH), luteinizing hormone (LH), total testosterone, fasting blood glucose, estradiol, thyroid-stimulating hormone (TSH), and dehydroepiandrosterone sulfate (DHEAS) were measured.

All participants were given a brief questionnaire to collect general information on demographics, health, medication use, lifestyle-eating habits, and illness conditions that may affect the gut microbiota and alter gut bacterial enzyme activity.

### Fecal specimen collection

After informed consent, participants were asked to collect fecal samples at home. They were supplied specimen collection kits, including a freezer pack and a barcoded container with exemplified instructions for collecting specimens. After fecal collection, the fresh samples were transported directly to our laboratories on pre-frozen freezer packs within two hours and stored at -80° C until analysis.

### Protein extraction

Protein extraction and enzymatic activities were carried out as described by (11) Flores R et al., 2012 with slight modifications to optimize the detection of the enzymatic activities. Approximately 0.5 gm of thawed feces in 5 ml of phosphate-buffered saline (PBS) was transferred to a 10 mL conical tube containing 5 mL of extraction buffer (60 mM Na2HPO4, 40 mM NaH2PO4, 10 mM Kcl, 1 mM MgSO4) and kept on ice. Fecal material was treated by heavy vortex for 1 min, and bacterial cells were lysed by sonication at maximum power for 90 s (30s intervals) on an ice bath. Lysates were centrifuged at 7000 rpm for 30 min at 4° C. The supernatant containing extracted proteins was transferred to new tubes and was used to measure protein concentration and enzymatic activities. Protein concentration in each lysate was estimated using the Folin Lowry method.

### Enzyme assay

Gut microbial enzyme activity was measured as follows: the reaction mixture (total volume of 0.5 ml) contained 0.25 ml of 2 mM p-nitrophenyl β-D-Glucuronide or/and 2mM p-nitrophenyl-β-D-glucopyranoside for β-D-glucuronidase and β-glucosidase respectively, and 0.25 ml of fecal suspension. The assay mixture was incubated at 37 c for 15 min for β-glucuronidase and 60 min for β-glucosidase. The reaction was quenched by adding 0.5 ml of 80mM of glycine buffer pH 10 and 0.5 N NaOH for β-glucuronidase and β-glucosidase, respectively, and vortex it. The absorbance of the product of both enzymes, i.e., p-nitrophenol, was measured at 540 nm and 420 nm for β-glucuronidase and β-glucosidase, respectively.

This method determines the amount of p-nitrophenol released after hydrolysis of p-nitrophenyl-D-glucuronide and p-nitrophenyl-D-glucopyranoside substrates. The enzyme’s activity can be calculated by comparing sample values to a standard curve prepared with p-nitrophenol. A unit of activity was accepted as such amount of p-nitrophenol for β-glucuronidase and β-glucosidase expressed in mM released during the reaction throughout 1 h per 1 mg of Protein. All data are mean and SD of at least three repeats.

### Statistical analysis

The software program GraphPad Prism 9 was used to process the experimental data. The data are shown as mean ± SD. All the experimental data were normally distributed or transformed to normal distribution. The statistical difference across the groups was assessed using a one-way ANOVA, t-test, and chi-square test, and p<0.05 was considered significant.

Pearson’s correlation was used to evaluate the relationship between the two enzymes’ activities. The relationship between the two enzymatic activity levels and Clinical parameters was estimated with Pearson’s correlation. Significance was based on trend or two-sided categorical tests with α = 0.05.

## Result

### Demographic, clinical, and biochemical parameters of the study subjects

The clinical characteristics of the study participants are summarized in Table 1. PCOS and the controls were matched for age and BMI. Compared to the control group, women with PCOS showed significantly higher levels of testosterone (p <0.05), estradiol (p <0.05), LH/FSH ratio (p <0.05), and hirsutism score (p <0.05). The levels of TSH, LH, prolactin, and DHEAS were not different among the groups.

**Table 1.** Demographic and clinical parameters of study participants

| Demographic variables and Clinical features |  |  |  |
| --- | --- | --- | --- |
| Variables & Parameters | Control group | PCOS group | *P* value |
| N | 25 | 23 | - |
| Age (year) | 27.47 ± 5.8 | 27 ± 6 | - |
| Body Mass Index (kg/ m<sup>2</sup>) | 24.76 ± 5.6 | 24.98 ± 5.15 | >0.05 |
| Waist-to-hip ratio | 0.77 ± 0.05 | 0.78 ± 0.07 | >0.05 |
| LH (mIU/ml) | 5.22 ± 3.1 | 6.30 ± 4.3 | >0.05 |
| FSH (mIU/ml) | 11.29 ± 7.8 | 7.99 ± 2.5 | **0.05** |
| LH/FSH | 0.55 ± 0.29 | 0.84 ± 0.73 | **0.04** |
| PRL (ng/ml) | 19.14 ± 9.74 | 26.84 ± 32.71 | >0.05 |
| T (ng/dl) | 14.67 ± 4.9 | 34.56 ± 36.08 | **0.01** |
| E2 (pg/ml) | 84.09 ± 18.04 | 69.38 ± 23.24 | **0.01** |
| TSH (uIU/ml) | 2.17 ± 1.4 | 1.8 ± 0.99 | >0.05 |
| DHEAS (ug/dl) | 143.56 ± 50.04 | 166. 17± 105.05 | >0.05 |
| Hirsutism (Ferriman-Gallwey) | 5.91 ± 2.8 | 9.65 ± 5.9 | **0.001** |
| Gut bacterial enzyme activity |  |  |  |
| Total Protein (mg/ml) | 1.35 ± 0.28 | 1.43 ± 0.31 | >0.05 |
| β-glucuronidase (mM/mg/hr) | 0.04 ± 0.01 | 0.05 ± 0.01 | **0.01** |
| β-glucosidase (mM/mg/hr) | 0.09 ± 0.05 | 0.13 ± 0.08 | >0.05 |

### Gut microbial β-Glucuronidase and β-Glucosidase enzyme activity differed between PCOS and Control Groups

To understand the relationship between the gut microbial enzyme activity and PCOS fluctuations, the fecal β-glucuronidase and β-glucosidase activities of the women with PCOS and healthy control were measured. To determine the activity of β-glucuronidase and β-glucosidase from the fecal specimens, we used the regression formula obtained from the calibration curve of p-nitrophenol (PNP), i.e., y = 7.9262x + 0.0085 (Supplementary 1). Table 1 shows the average level of β-glucuronidase activity and β-glucosidase in PCOS and healthy control stool specimens. β-glucuronidase activity differed statistically significantly between the PCOS group and the controls (0.05±0.01 vs. 0.04±0.01, p=0.01) (figure 3). In contrast, β-glucosidase activity did not differ significantly between groups but was higher in the PCOS group (0.13±0.08 vs. 0.09±0.05, p=0.06) (figure 3).

**Figure 1.**
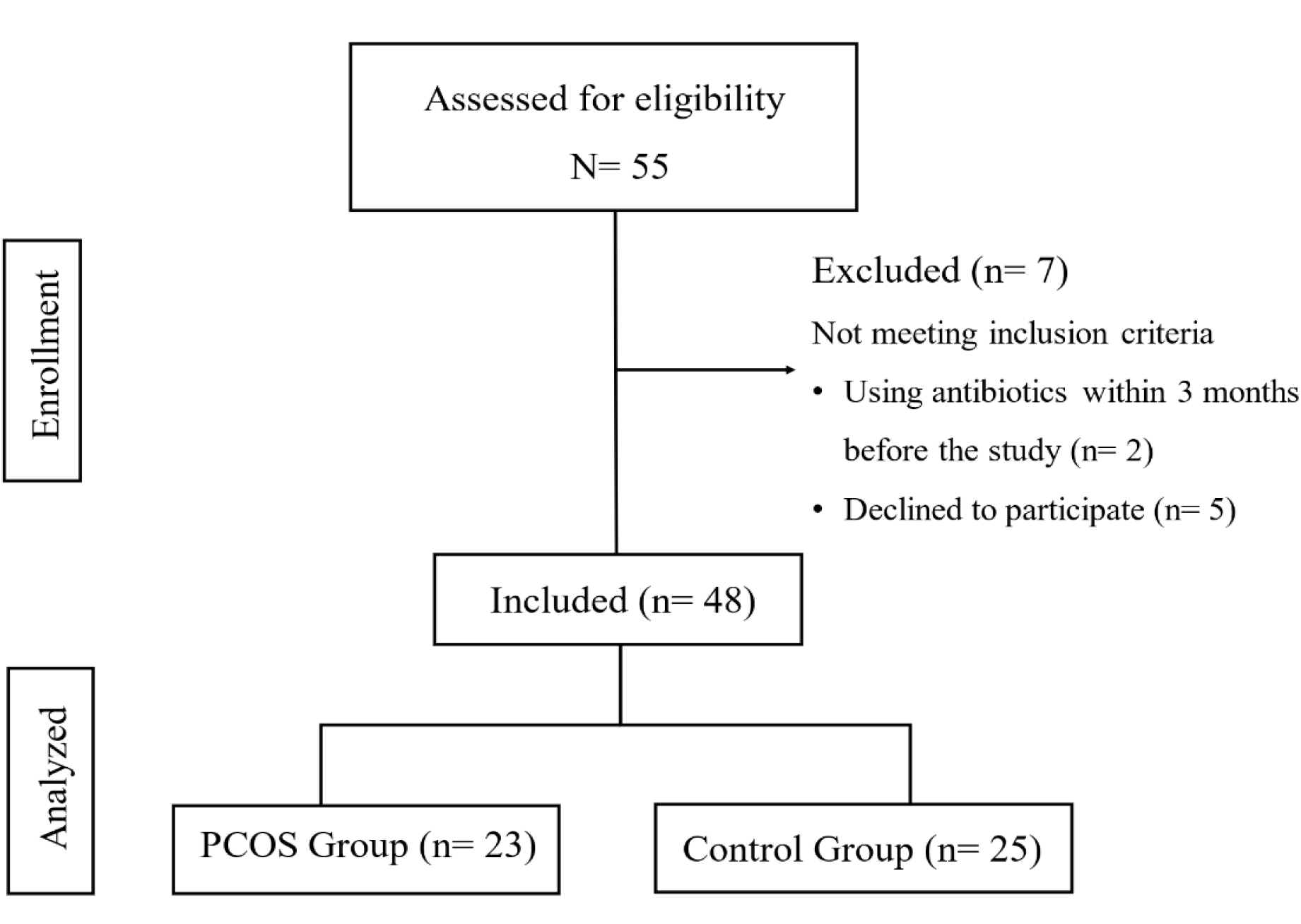
Flow chart showing participants recruitment in the study.

**Figure 2.**
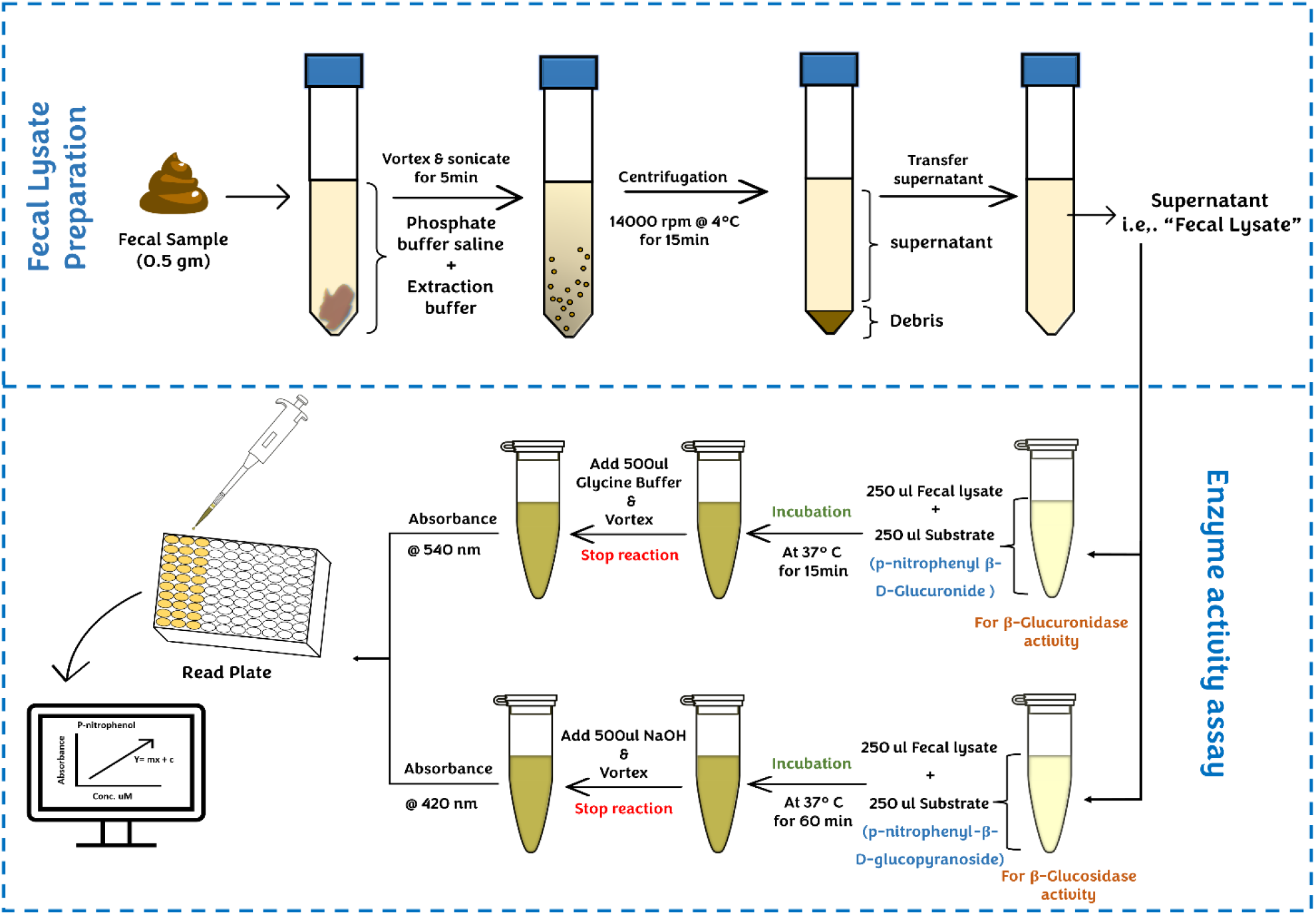
Method overview of fecal lysate preparation and enzyme activity assay. Fresh-frozen or freshly collected fecal pellets were weighed, followed by vortex and sonicated in a mixture of phosphate buffer saline pH 7.4 and extraction buffer. Following centrifugation, the supernatant was isolated (fecal lysate) and used in a colorimetric enzymatic assay. The fecal lysate was divided into two tubes, and an enzyme assay was done for β-glucuronidase and β-glucosidase.

**Figure 3.**
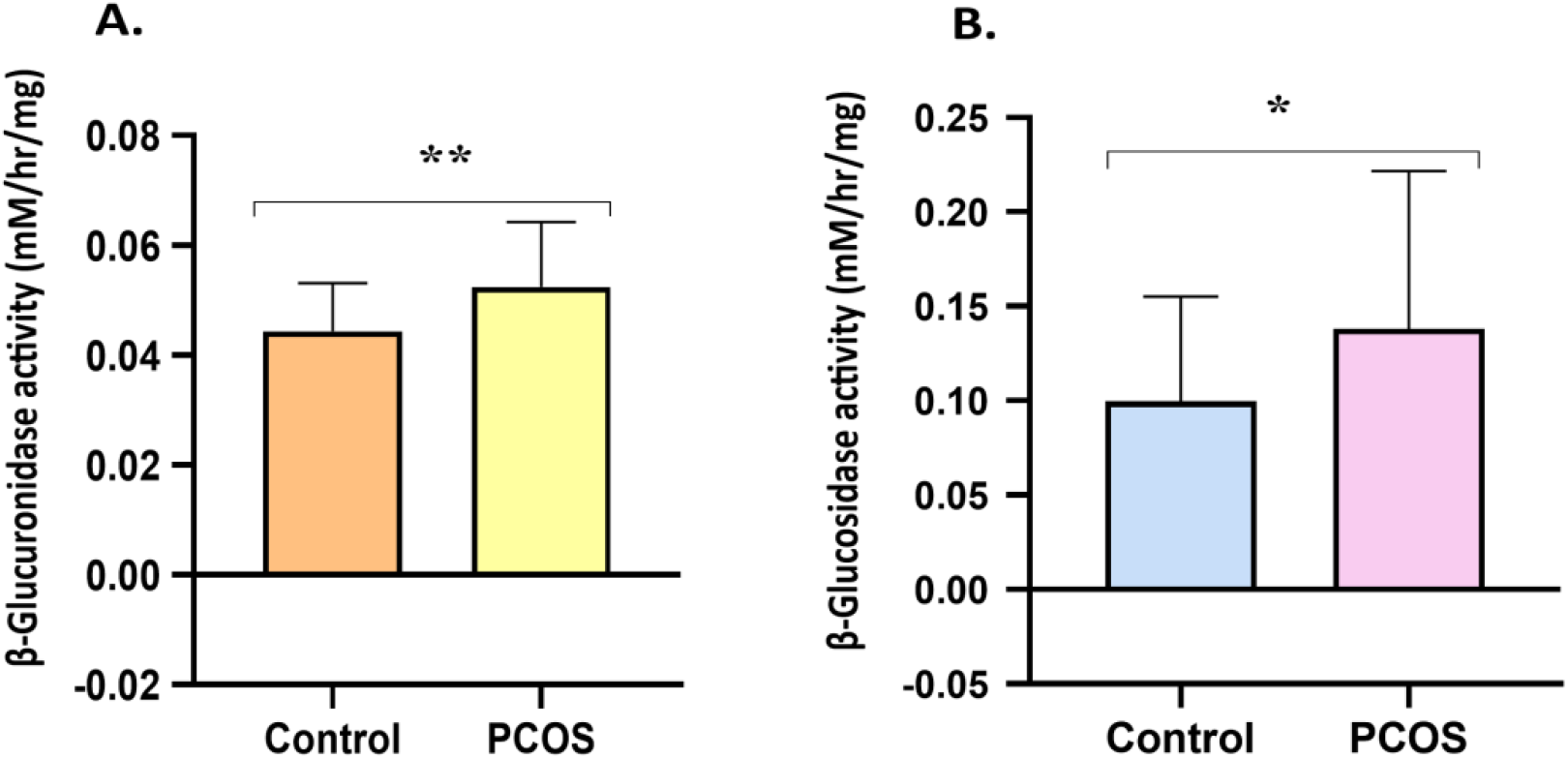
Average activity of Gut microbial enzymes. **A**. PCOS group present a higher level of gut microbial β-glucuronidase activity compared to Control groups with p <0.05 **B**. Gut microbial β-glucosidase activity shows high in the PCOS group compared to control groups with p >0.05.

### Correlation between β-Glucuronidase and β-Glucosidase activity

We wondered if the enzymes’ activities were correlated. Therefore, we performed a Pearson correlation analysis of β-Glucuronidase activity and β-Glucosidase activity in each group. Interestingly, we found a statistically significant positive correlation between β-Glucuronidase activity and β-Glucosidase activity in the PCOS group (r =0.55; p=0.005 (figure 3(A)), Whereas in the controls, we found a negative correlation between β-Glucuronidase activity and β-Glucosidase activity (r =-0.28; p=0.16) (figure 4 (B)), although it was not statistically significant.

**Figure 4.**
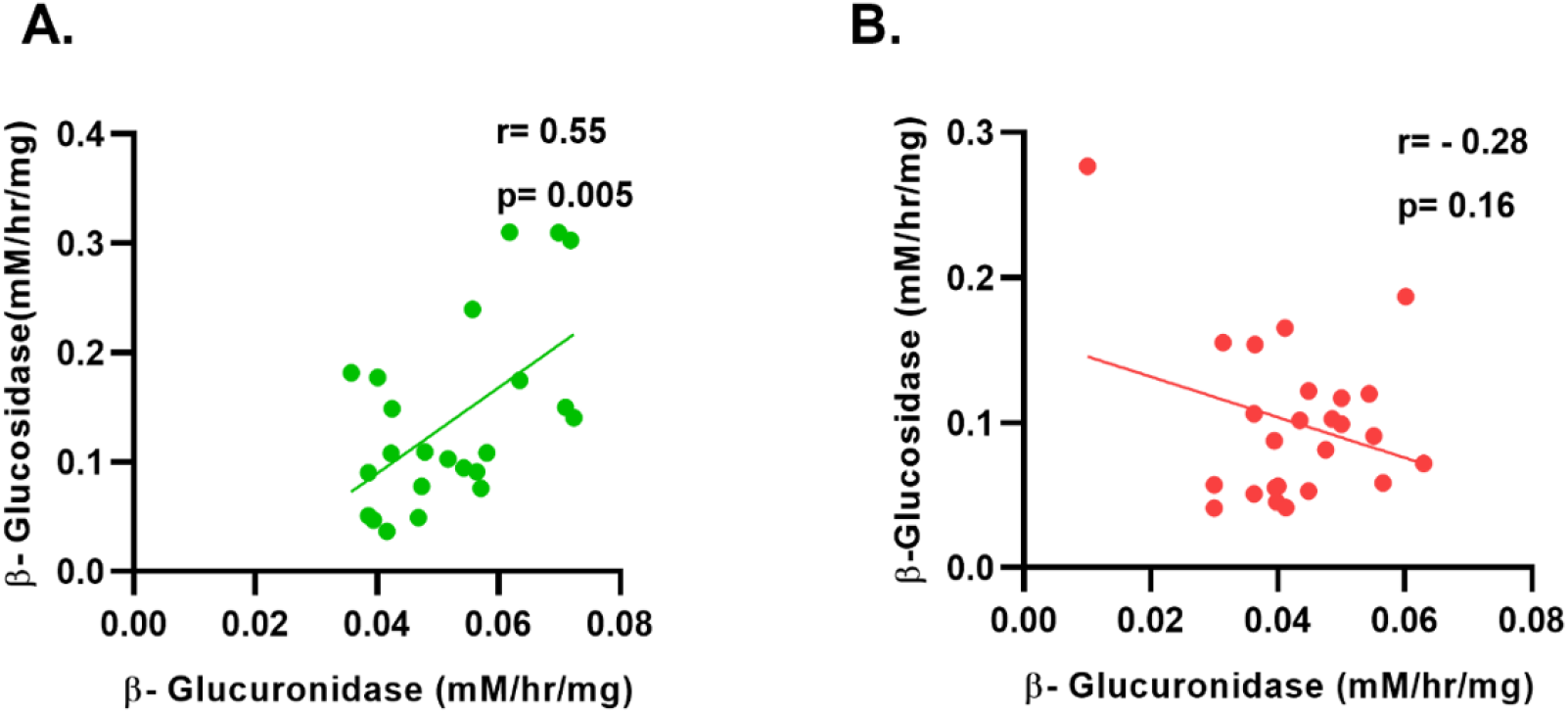
Correlation analysis of gut microbial β-glucuronidase and β-glucosidase activities. **A**. PCOS group shows a positive association between β-glucuronidase and β-glucosidase activities with r=0.55 and p=0.005 **B**. The control group shows a negative association between β-glucuronidase and β-glucosidase activities with r=0.16 and p=0.16.

### Correlation between gut microbial enzyme activity and serum sex hormone levels

According to the Pearson correlation analysis of β-Glucuronidase activity with testosterone of PCOS and control groups, a moderate positive correlation existed in the PCOS while a strong positive association was found in the control group (r= 0.10, p=0.64 vs. r=0.37, p=0.06 respectively) (fig. 5(C)(D)). However, it did not reach to statistical significance value. Wise, a positive correlation existed (r=0.3, p=0.17 vs. r=0.05, p=0.80 respectively) (fig. 5 (A)(B)) in the PCOS group and no association was found in control with estradiol which is also statistically insignificant.

**Figure 5.**
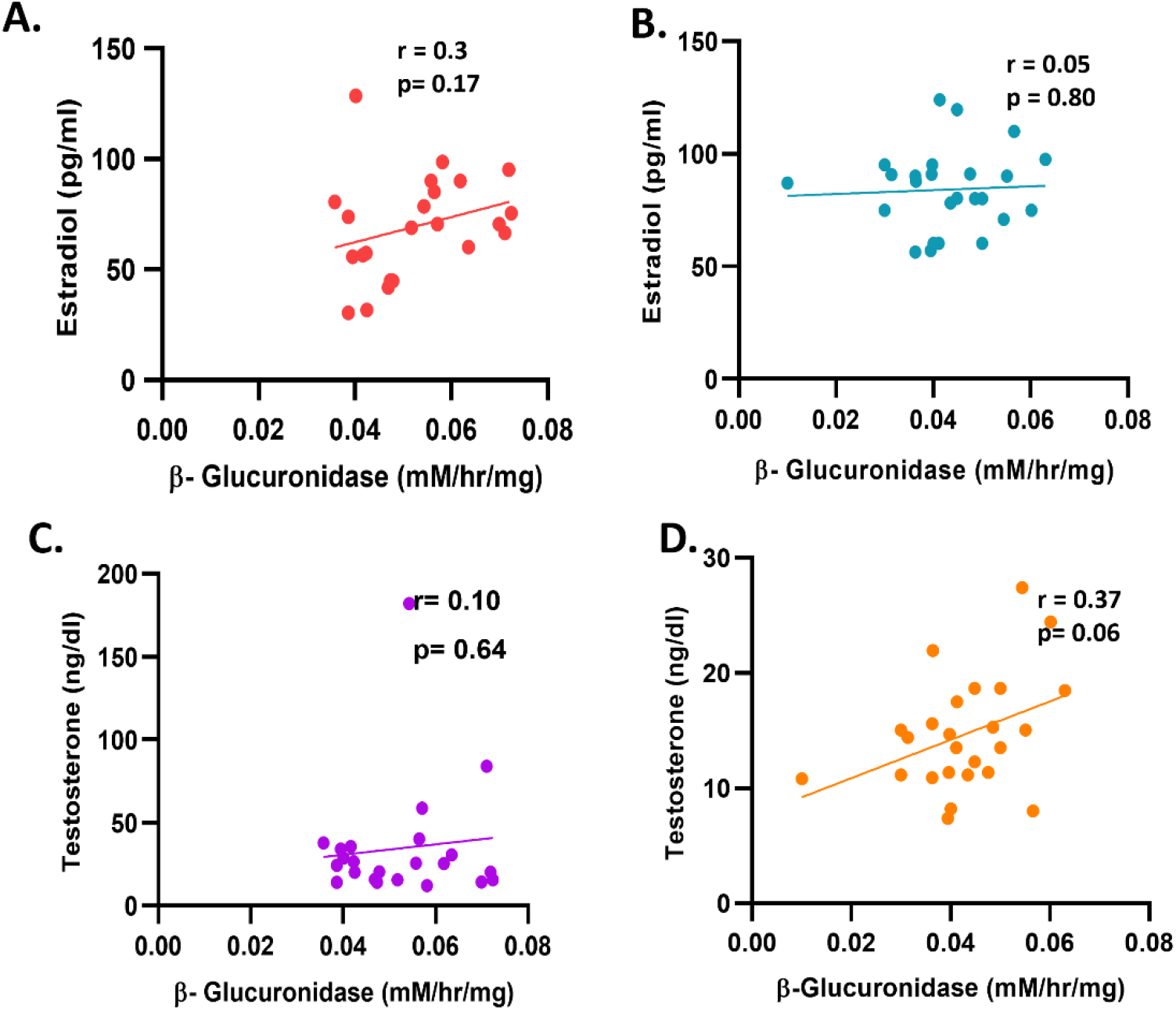
The correlation matrix of gut microbial β-glucuronidase activity with estradiol and testosterone levels **A**. PCOS group shows a positive correlation between β-glucuronidase activity and estradiol with r=0.3 and p=0.17 **B**. The control group shows a negligible positive correlation between β-glucuronidase activity and estradiol level with r=0.05 and p=0.80 **C**. PCOS group shows a positive association between testosterone level and β-glucuronidase activity with r=0.10 and p=0.64 **D**. Control group shows a positive association between testosterone and β-glucuronidase activity with r=0.37 and p=0.06.

We also examined the relationship between β-glucosidase activity with testosterone and estradiol in PCOS and the control group. Here we found a statistically significant positive correlation between β-glucosidase activity and estradiol in the PCOS group (r=0.48, p=0.01) (figure 6 (A)). In contrast, in the control group, we found a contradictory result of a negative correlation (r=-0.25, p=0.22) (figure 6 (C)), but it was not statistically significant. In the case of the association between β-glucosidase activity and testosterone, the study found a negative correlation in the PCOS group (r=-0.14, p=0.52) (figure 6 (B)). In contrast, a positive correlation existed in controls (r=0.17, p=0.39) (figure 6 (D)), although it did not reach to statistical significance value in the groups.

**Figure 6.**
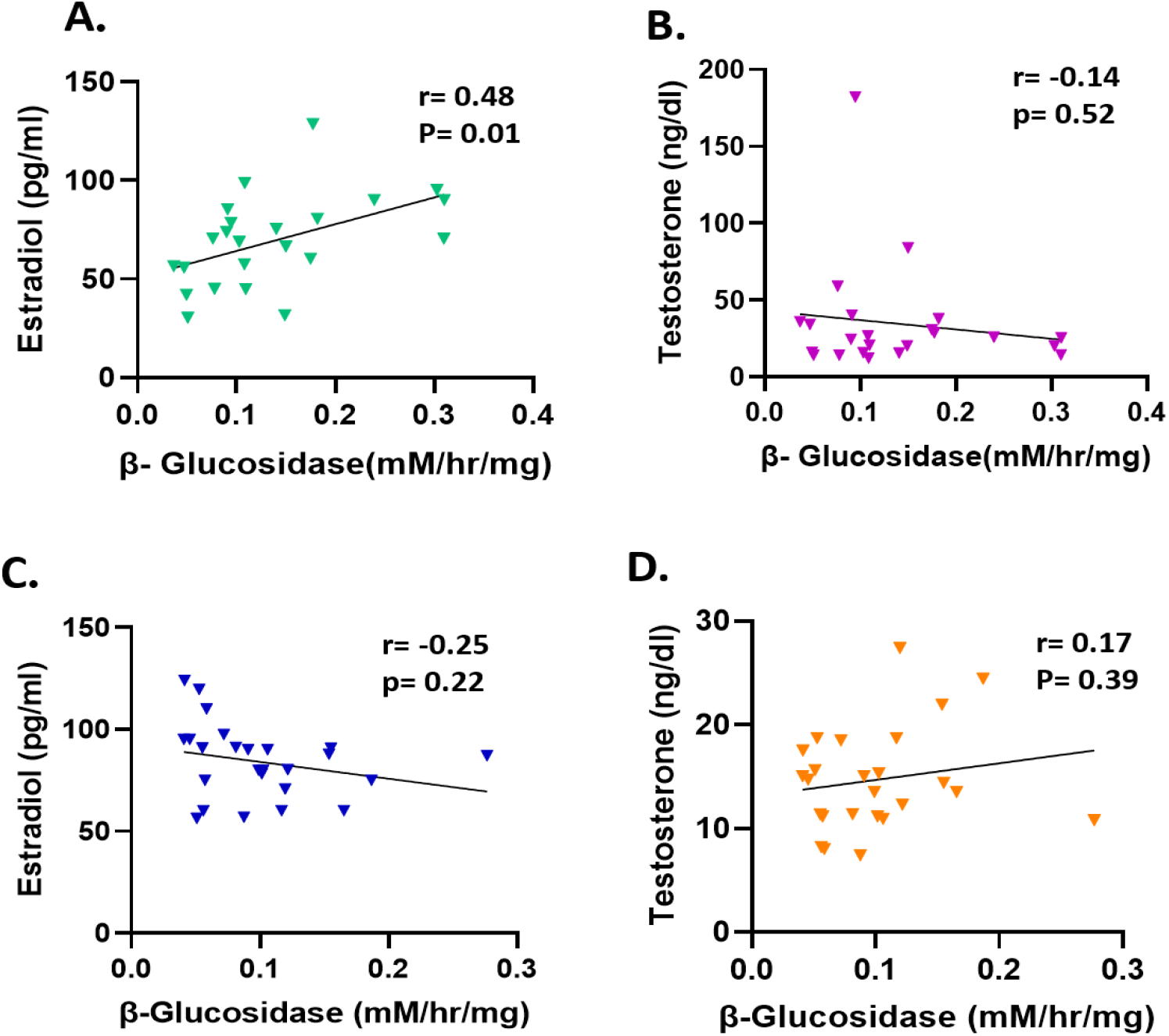
Correlation matrix of gut microbial β-glucosidase activity with estradiol and testosterone level **A**. PCOS group shows a positive correlation between β-glucosidase activity and estradiol with r=0.48 and p=0.01 **B**. PCOS group shows a negative correlation between β-glucosidase activity and testosterone level r=-0.14 and p=0.52 **C**. The control group shows a negative association between estradiol level and β-glucosidase activity with r=-0.25 and p=0.22 **D**. Control group shows a positive association between testosterone and β-glucosidase activity with r=0.17 and p=0.39.

## Discussion

Our study explored the distinctions in gut microbial β-glucuronidase and β-glucosidase activities between women with PCOS and those without. By doing so, we hoped to shed new light on these enzymes’ role in this condition.

This investigation provided new insights into the differences in gut microbial β-glucuronidase and β-glucosidase activities between women with PCOS and controls. While previous research by Karolina et al. (2022) only explored the relationship between β-glucuronidase and androgen levels in overweight and obese women with PCOS, our investigation delved deeper, revealing that both β-glucuronidase and β-glucosidase activity levels were significantly higher in women with PCOS, especially β-glucuronidase, which was statistically significant (12).

Our study also revealed intriguing connections between hormone levels and the activity of these enzymes. Specifically, testosterone was positively correlated with β-glucuronidase activity in both groups. As a rodent study had shown, gut bacteria-driven deglucuronidation of androgens significantly affected androgen metabolism, suggesting that this enzyme could impact systemic testosterone levels (13). Additionally, we found a moderate negative relationship between β-glucosidase activity and testosterone in women with PCOS, contrasting with a strong positive association in the control group. However, the functional significance of these findings remains unexplained.

In keeping with the literature (14), the study found that women with PCOS had lower levels of estradiol and much higher levels of testosterone than the control group. Moreover, estradiol was positively associated with β-glucuronidase activities in both groups, although the correlation was not statistically significant. This chimes with the idea that β-glucuronidase allows estrogen to re-enter circulation, increasing the estrogen pool, and that a rise in β-glucuronidase-producing bacteria could elevate circulating estrogen levels. Furthermore, we discovered a significant positive correlation between β-glucosidase activity and estradiol in the PCOS group, while a negative correlation was found in the control group. This indicated a complex interplay between hormonal factors, particularly estradiol, and the activities of β-glucuronidase and β-glucosidase.

In conclusion, our study uncovered crucial differences in gut microbial enzyme activity among women with PCOS in the urban region of Gujarat, revealing a multifaceted relationship between enzyme activity and hormones. This study raises a possibility that by monitoring gut microbial enzyme activity, produced by specific bacterial species, we could better understand and potentially modulate microbial dysbiosis in the clinical management of PCOS. Further research into the gut microbiome of obese women, both with and without PCOS, could help clarify the role of obesity and insulin resistance in shaping gut microbial activity in PCOS. However, given the vast diversity of the human gut microbiome, extensive clinical cohorts were necessary to address these questions comprehensively.

Our study has limitations, including a small sample size and a cross-sectional design. Additionally, the participants in both the PCOS and control groups were matched based on BMI, which could overlook other relevant factors. The study also lacked expression data or in vitro systems to investigate the regulatory mechanisms and factors influencing microbial enzymatic activity. Furthermore, it is essential to consider that metagenome analysis might have provided a deeper understanding of the alterations in gut enzyme activities in PCOS. Despite these limitations, our study offers valuable insights into the differences in gut microbial enzyme activities in women with and without PCOS.

## Supporting information

SL Fig. 1 Calibration curve of p-nitrophenol

## Data Availability

The datasets used and analyzed during the current study are available from the corresponding author upon reasonable request.

## Abbreviations

PCOS: Polycystic ovary syndrome
NAFLD: non-alcoholic fatty liver disease
BMI: body mass index
WHR: waist to hip ratio
FSH: follicle stimulating hormone
LH: luteinizing hormone
TSH: thyroid stimulating hormone
DHEAS: dehydroepiandrosterone sulfate
PRL: prolactin
E2: estradiol
T: testosterone
PNP: p-nitrophenol
GM: gut metabolite
gmGUS: gut microbial β-glucuronidase.

## Acknowledgments

We thank all the study participants and Gujarat University health center clinicians for their assistance. Furthermore, we are deeply grateful to the Scheme of Developing High-Quality Research (SHODH) Department of Education, Government of Gujarat, India, for providing fellowship to Jalpa Patel and CSIR-UGC-NET, providing companionship to Hiral Chaudhary.

## Authors’ contributions

JP assisted with data collecting, writing, and manuscript preparation. HC assisted with data collection. RJ and BP drafted and revised this manuscript. RJ and BP carried out the critical review.

## Authors approval

All authors approved the submitted version and agreed to be personally accountable for their contributions and ensure that questions related to the accuracy or integrity of the work are appropriately investigated, and the resolution is documented in the literature.

## Funding

No funding was involved with this study.

## Ethics approval and consent to participate

The Institutional Ethics Committee (IEC) of the University School of Sciences, Gujarat University, approved the study.

## Competing interests

The authors declare that they have no competing interests.

## Notes

### Competing Interest Statement

The authors have declared no competing interest.

### Funding Statement

This study did not receive any funding.

### Author Declarations

The Institutional Ethics Committee (IEC) of the University School of Sciences, Gujarat University Ref. Letter No. :GU-IEC(NIV)/02/Ph.D./007

