## Supplementary figures and images for "Assessment of Gut Microbial β-Glucuronidase and β-Glucosidase Activity in Women with Polycystic Ovary Syndrome"

### SL Fig. 1 Calibration curve of p-nitrophenol

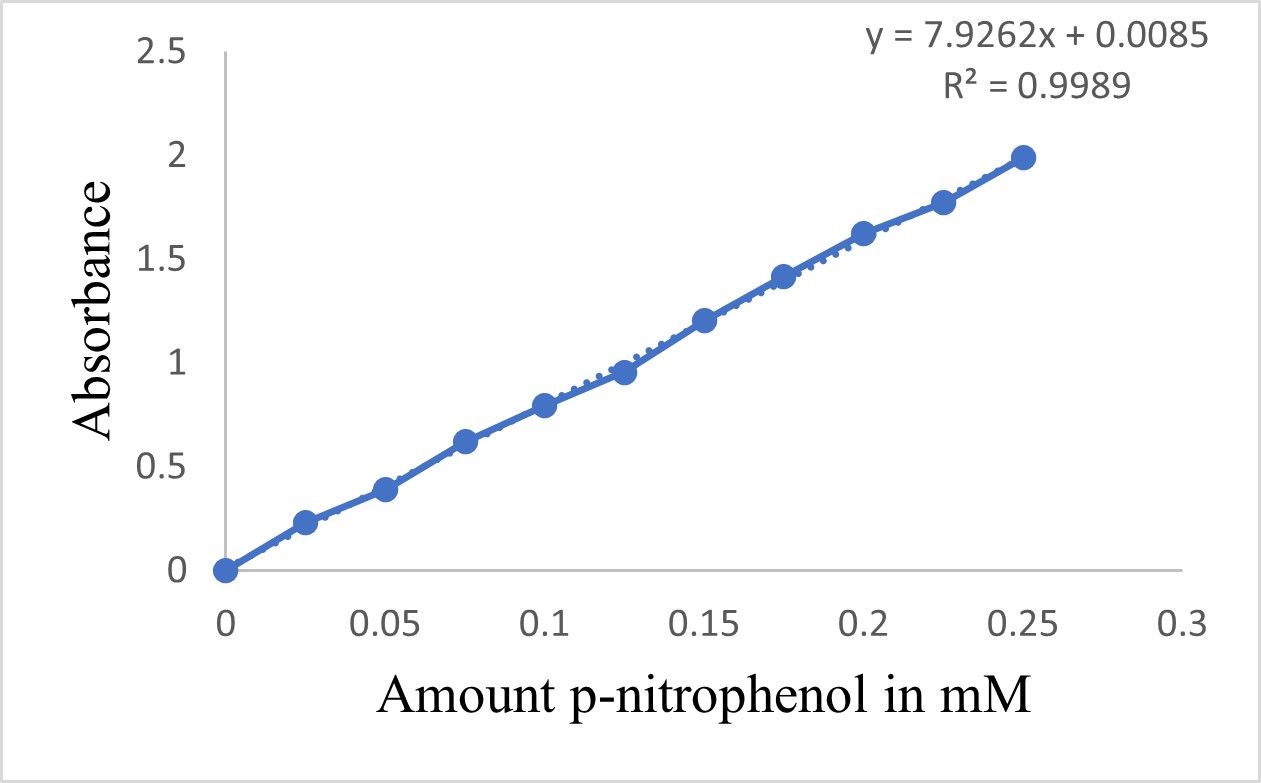
